# Presbyopia onset affects dynamic visual acuity via motor adaptation in naturalistic viewing conditions

**DOI:** 10.1101/2025.04.04.25325235

**Authors:** Mira Baroud, Hadrien Jan, Martha Hernandez, Stephane Boutinon, Denis Sheynikhovich, Delphine Bernardin

**Affiliations:** Sorbonne Université, Inserm, CNRS, Institut de la Vision, F-75012 Paris, France; Center of Innovation and Technologies Europe, Essilor International, SAS, Charenton-le-Pont, France

**Keywords:** dynamic visual acuity, aging, presbyopia, motor coordination, vergence, accommodation

## Abstract

This study investigated how age affects dynamic visual acuity (DVA) and its visuomotor correlates during a high-acuity real-world visual task, focusing on the roles of motor coordination and binocular mechanisms around the onset of presbyopia. Sixty-three participants, ranging from young and adult non-presbyopes to presbyopia-onset individuals and older presbyopes, performed a smartphone-based DVA test while sitting, standing, and walking. The participants’ visuomotor profile was assessed by 8 vergence and accommodation metrics along with 8 motor variables related to smartphone positioning relative to the head. Findings revealed a significant age-related decline in DVA, with the sharpest drop occurring at presbyopia onset. This decline was closely linked to reduced visual accommodation, while vergence had minimal influence. Motor variables, particularly smartphone distance and its antero-posterior and lateral movements, showed a stronger overall impact on DVA. The results suggest that diminished motor adaptation and age-related accommodative changes jointly contribute to the deterioration of DVA during presbyopia onset.

## 1. Introduction

Mobility is fundamental to normal aging and is intimately linked to health status and quality of life (Webber et al., 2010). The visual component of mobility is tested by the dynamic visual acuity (DVA), defined as the capacity to discern fine details of a visual stimulus when there is relative motion between the observer and the stimulus (Long & Penn, 1987). A strong and progressive age-related decline in DVA was observed in studies with computerized tests in seated participants (Li et al., 2014) and in participants walking on a treadmill while observing visual stimuli fixed to the wall or to the head (Borg et al., 2015; Hirasaki et al., 1999; Verbecque et al., 2018). However, it is not clear whether this decline reflects functional deficits in standard real-world viewing conditions, where multiple sensory-motor adaptation mechanisms help coordinate body, head and eye movements to stabilize the image and maintain clear vision (Imai et al., 2001) as well as balance while in motion (Imai et al., 2001; Kavanagh et al., 2005; Long & Penn, 1987; Webber et al., 2010).

While not used in previous DVA studies, consulting a mobile device during walking is a highly realistic dynamic high-acuity visual scenario, relying on both visual and motor mechanisms of retinal image stabilization. It is also highly relevant for life quality, since mobile devices have become an essential tool in people’s daily life with 7 billion users worldwide by the end of 2024, 72% of which are over 65 years old (Statista Research Department, 2025; Wang et al., 2025). Recent studies on smartphone holding behavior report that 74.3% of users continue walking when consulting it and adopt a so-called “block motor” strategy, in which the arms and head move in phase with the thorax to reduce the relative motion of the phone (Argin et al., 2020; Schabrun et al., 2014). These findings suggest an important role of motor adaptation. In addition, binocular visual mechanisms related to accommodation, vergence and their coupling (Miles et al., 1987) are likely to play a role. Indeed, normal aging leads to a decline in both vergence- and accommodation-related measures (Anderson et al., 2008; Heron et al., 2001; Mordi & Ciuffreda, 2004; Rambold et al., 2006; Yekta et al., 1989) with some of these measures strongly affected by presbyopia onset. The relative contribution of motor adaptation and binocular mechanisms in DVA has not been studied so far.

This study addresses these gaps by measuring DVA using a smartphone application in adult subjects just before and during the presbyopia onset period, in addition to younger (non-presbyopic) and older (mature presbyopic) participant groups used in previous studies. To test the role of binocular mechanisms in age-related DVA changes, the latter are correlated with a number of accommodation and vergence variables. To test the effect of motor adaptation, smartphone position in the head-referenced frame was recorded, providing distance and orientation information as well as total amount of smartphone movement.

## 2. Methods

### 2.1. Participants

A total of 63 participants were recruited. They were divided into four age groups: 17 young non-presbyopic group (YNP, 7 females, 18-25 yo, μ= 21,35 ± 2,11), 16 adult non-presbyopic group (ANP, 8 females, 28-35 yo, μ =30,72 ± 2,19), 15 presbyopia onset group (PO, 11 females, 40-46 yo, μ=43 ± 2) and 15 mature presbyopic group (MP, 9 females, 51-65 yo, μ=58,53 ± 5,09). Among the YNP participants, 4 (2 myopic and 2 hyperopic) wore single-vision habitual corrective lenses. Among the ANP participants, 11 had myopia and wore single-vision corrective lenses. Among the PO participants, 1 wore a progressive addition lens (PAL), and 8 wore single-vision lenses. Finally, all the participants in the MP group wore PAL. All participants reported satisfaction with their optical equipment. Included participants reported no visual, cognitive or sensorimotor pathology, deficit, or disorder, apart from normal age-related visual changes. All participants had visual acuity at far and near distances ≧20/25 (0.1 logMAR) and stereoscopic acuity of at least 50 arc-min as measured by Wirt test. Before participating in the study, all participants gave informed written consent. Recruitment and all experimental procedures were in accordance with the tenets of the Declaration of Helsinki and they have been approved by the île-de-France VII Personal Protection Committee (number SI: 23.00499.000186).

### 2.2. Equipment

All behavioral variables were measured using iPhone 12 (Apple Inc. Cupertino, California, US) equipped with IOS 17.1.2. An integrated Inertial Measurement Unit (IMU) measured phone’s orientation in space at the sampling frequency of 100 Hz. An embedded TrueDepth camera (30 Hz) was used to measure the position of the smartphone with respect to the user head. 3D coordinates of several predetermined points of the user face were recorded in the head reference frame (positive X, Y, Z directions correspond to the egocentric left, up, and towards the viewer, respectively).

### 2.3. Visual stimuli and acuity measurement

Landolt C optotype was presented on the smartphone’s screen in one of 8 possible orientations. Participants identified the orientation of the optotype by pressing a button. A 15-step staircase Bayesian procedure was used to determine visual acuity (Piech et al., 2020). Two stimulus presentation modes were used during DVA tests: (i) In the *active visual acuity* (AVA) mode, the size of the stimulus on the screen was automatically updated depending on the distance between the screen and the observer (i.e. the optotype occupied an equivalent space in the field of vision independently of the smartphone distance); (ii) In the *behavioral visual acuity* (BVA) mode, the visual size of the optotype did not adapt to the smartphone distance and maintained a fixed size on the screen. While the BVA mode is more natural, the AVA mode permits the evaluation of visual acuity in the absence of accommodation-vergence cross-coupling and of the magnification visual cues of the optotype. The time delay between the change in distance and the update of the stimulus size on the screen (in the AVA mode) was <20 ms ensuring smooth visual experience during acuity measurement tasks.

### 2.4. Protocol

DVA was measured in three different conditions. In the *sitting condition*, participants sat on a chair, without a table, with their feet flat on the floor. In the *standing condition*, they stood with their feet hip-width apart. Finally, in the *walking condition*, participants walked at their habitual speed back-and-forth along an L-shaped hallway (total length: 8 m) during the time period of the DVA estimation. In all conditions, the participants were instructed to hold the smartphone as they usually do in daily life and were free to adopt their body posture, gait and smartphone holding position. In total, each participant performed 3 conditions in 2 visual acuity measurement modes with 2 repetitions each in randomized order. Before conducting the experiment, there was an unrecorded familiarization trial and static visual acuity (SVA) was measured while the participants were asked to put both hands on the table to maintain stability and to hold the smartphone at a fixed distance of 30 cm. On average, the experiment lasted about 1 hour per participant.

### 2.5. Refraction, accommodation and vergence measures

Before the DVA tests were conducted, refraction, accommodation and vergence functions were assessed by a certified optometrist. All measures are summarized in Table 1.

**Table 1.** Refraction, accommodation and vergence measures collected in all participants. From the subjective refraction variables, only the first one (sphere) was used in regression analyses.

| Variable | Method and/or Equipment | Distance |
| --- | --- | --- |
| Subjective refraction: sphere, cylinder, axis in OD, OS | Vision-R™ 800, EssilorLuxottica | 5m |
| Near point of convergence (NPC, cm) | Distance corresponding to the maximum effort of the convergence | - |
| Associated phoria (-EXO, + ESO) | Mallett test | 40cm |
| Sheard's Ratio | Between dissociated phoria and fusional vergence reserve | - |
| Accommodative-convergence over accommodation (AC/A) ratio | Vision-R™ 800, +1 lens | 40cm |
| Near point of accommodation (NPA, cm) | Distance corresponding to the maximum accommodative power | - |
| Accommodative facility (cycles/minute) | +/- 2,00 for YNP and ANP<br>+/- 1,00 for OP | 40cm |

### 2.6. Behavioral measures

From the timeseries recordings of the smartphone distance (between the phone and the head, in cm), the pitch, yaw and roll angles (in degrees of visual field in the YZ, XZ and XY planes, respectively) by the TrueDepth camera, the following 8 variables were calculated: walking pace; average distance pitch, yaw and roll angles, computed between the timestamps of optotype presentation and a participant’s response; average time derivatives of the distance, pitch and yaw data over the same period (henceforth termed antero-posterior movement, vertical movement, lateral movement).

To assess the change in smartphone holding distance depending on the task difficulty, the *relative distance* was calculated by the difference between the 2nd step (large visual stimulus and hence low task difficulty) and the last step (small stimulus and high task difficulty) of the staircase VA procedure (the first step was discarded due to high transient variability associated with walking initiation).

### 2.7. Data analysis

The collected data were processed using Matlab (R2023a, update 6) and the final dataset is available at the Mendeley data repository: https://doi.org/10.17632/dkswf7gzgm.1. Statistical analyses were performed using R software (version 4.5.0, R Core Team 2025). Mixed model ANOVAs with age group as a between-subjects factor and other variables as within-subjects factors were performed using R package rstatix (version 0.7.2). Homoscedasticity, and normality were verified visually through residual plots and quantitatively using the Shapiro-Wilk test. The effect size was determined by partial η^2^. Holm-Bonferroni corrections were used for post hoc comparisons. Where specified, data were analyzed using multiple linear regression to examine relationships between variables. For correlation analyses, when normality was not verified, Box-Cox transformations were applied to the variables. If transformed variables did not verify normality, Spearman’s correlation was computed instead of Pearson’s.

## 3. Results

### 3.1. Effect of age and postural condition on DVA

We first assessed the relationship between the two different DVA measures (i.e. AVA and BVA) in the novel smartphone-based paradigm (Fig. 1A). The two measures were highly correlated (r=0.84, p<10^-5^, R^2^=0.7) suggesting that such an approach provides a consistent measure of visual performance. To reduce measurement variability, we evaluated DVA by the average of AVA and BVA in the subsequent analyses. To compare the dependence of this DVA measure on age with previous visual acuity data in static (Elliott et al., 1995) and dynamic (Li et al., 2014) conditions, these data were plotted as a function of decade of age (Fig. 1B). While there is an obvious reduction of visual acuity due to movement, the availability of motor and binocular adaptation mechanisms in the smartphone-based paradigm leads to a striking increase in performance, compared to screen-based tests in seated participants. Moreover, while the results of the screen-based tests suggest a stable DVA until the age of 50, in our real-world paradigm the reduction in DVA was associated with the age of 40 near the presbyopia onset period.

**Figure 1.**
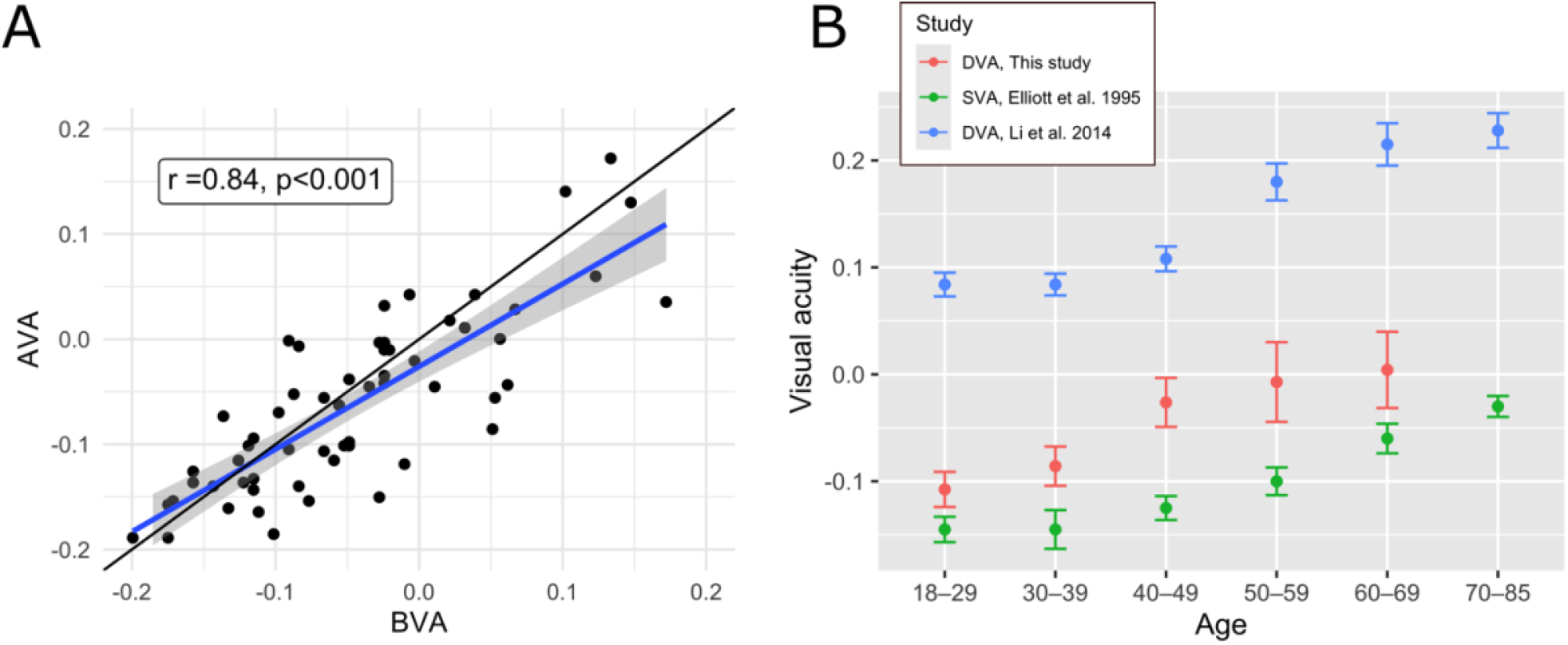
**A**. Relationship between AVA and BVA measures of DVA (see Methods) across the complete sample of subjects. Each dot corresponds to a participant. The blue line corresponds to linear regression (±*SE*) and the black line represents the unity relationship. **B**. Comparison of DVA measured in this smartphone-based study (in red), static visual acuity data by Elliott et al (1995) and the DVA measured in a computerized test in seated participants (Li et al., 2014). Visual acuity values are in logMAR.

This was further analyses by a two-way mixed ANOVA with the age group as an independent factor and the dynamic condition as a repeated measure (Fig. 2A). The main effect of age was significant (F_3,54_=6.756; p <.001; η^2^ =.27) showing that sensory-motor and binocular adaptation mechanisms do not fully compensate for DVA loss in aging. Multiple comparisons revealed a significant difference between ANP and PO participants (p=.008), but no significant differences between non-presbyopes (YNP vs ANP, p=.42) and between presbyopes (PO vs MP, p=.42), supporting the observation that presbyopia onset is the critical time period in DVA decline in realistic viewing conditions. The average difference between ANP and PO groups amounted to 0.06 logMAR corresponded to 3 letters the ETDRS chart.

**Figure 2.**
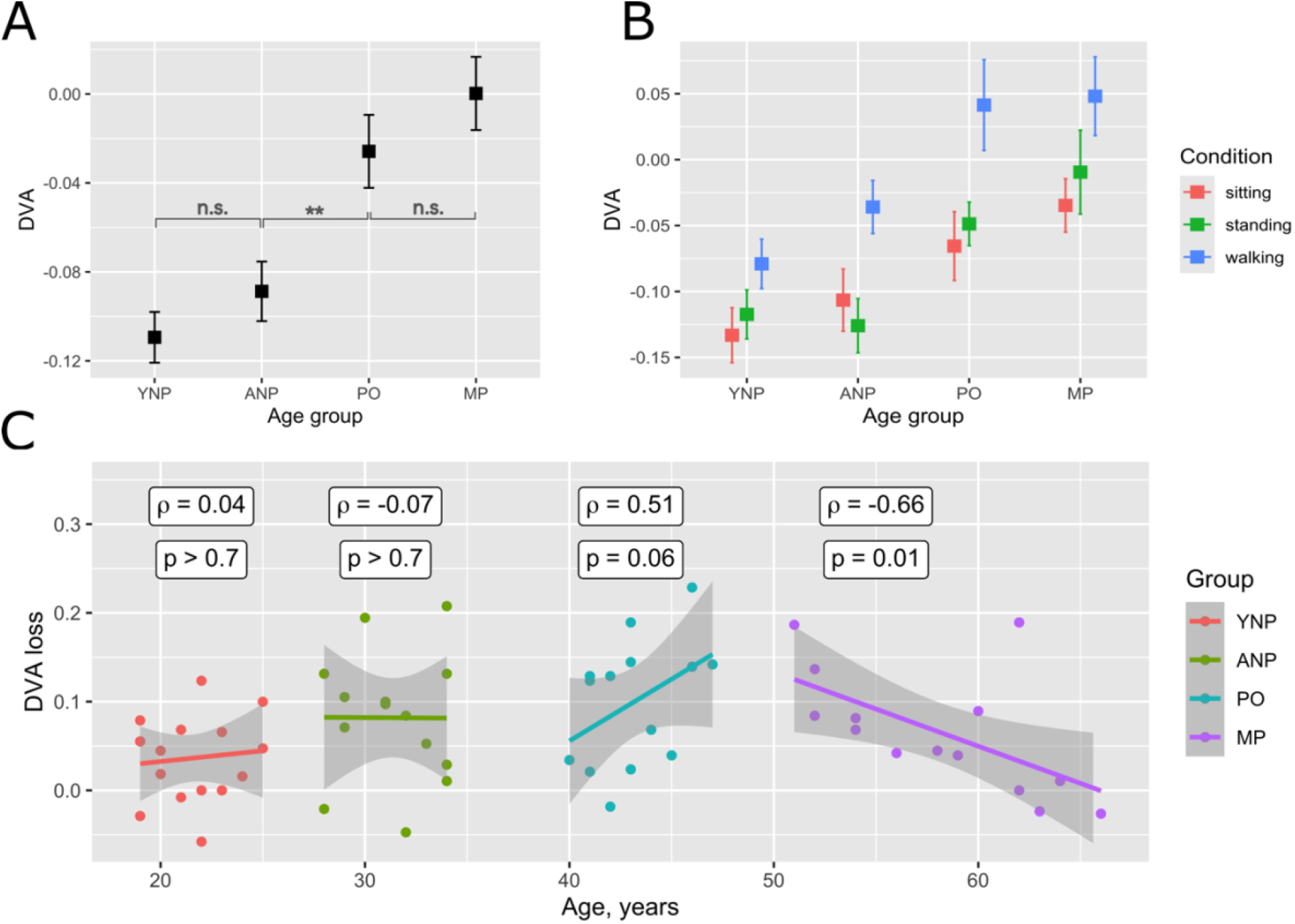
Impact of age and postural conditions on DVA. **A**. Effect of age on DVA. Black squares indicate mean values with error bars representing the standard error. **B**. Effect of postural condition (sitting, standing and walking) on age-dependent DVA. **C**. DVA loss (in logMAR) as a function of chronological age in the four subject groups. Linear regression fits (±*SE*) are shown by colored lines. YNP: young non-presbyopia group; ANP: adult non-presbyopia group; PO: presbyopia-onset group and MP: mature presbyopia group. Significance: ** (p<0.01), n. s. (p>0.05).

In the smartphone-based paradigm, DVA is likely to be affected by postural adaptations, with different complexity levels depending on the movement type (i.e. sitting, standing or walking). Accordingly, there was a significant main effect of postural condition (F_2,108_=33.896; p<.001; η^2^=0.39), explained by a strong degradation of DVA in the walking condition compared to the sitting and standing (Fig. 2B, walking – standing: p<10^-7^, walking – sitting: p<10^-8^, standing – sitting: p=0.45). The statistical interaction between age and postural condition was also significant (F_6,108_=2.209; p=0.048, η^2^=0.11), explained by the fact that in ANP and PO groups DVA during standing differed significantly from that during walking (ANP: p<0.001, YNP: p=0.008), whereas in the YNP and MP these differences were not significant. Walking vs sitting conditions differed significantly in all age groups tested, with middle-age groups (ANP and PO) experiencing the strongest decline in performance.

To further investigate these age-related differences, DVA loss associated with the switch from static to dynamic activities was measured by the difference between DVA during walking and the average over sitting and standing conditions. Whereas one-way ANOVA performed on the DVA loss across age groups did not reach statistical significance (F_3,54_=2.517; p=.068), we reasoned that more information can be obtained by using age as a quantitative variable rather than a factor (Fig. 2C). Correlation analyses revealed that in both non-presbyopic groups DVA was uncorrelated with age (YNP: Spearman’s ρ=0.04, ANP: ρ =-.07, p>0.7). In contrast, there was a statistical tendency for a positive correlation in the PO group (ρ =0.51, p=0.06) and a significant negative correlation in the MP group (ρ =-0.66, p=0.01), suggesting a greater DVA loss during presbyopia onset, which progressively diminishes with further aging.

### 3.2. Binocular contributions to DVA decline

We next assessed whether age-related changes in DVA (response variable) can be statistically accounted for by an age-related decline in vergence and accommodation mechanisms, in addition to chronological age and SVA. Out of all visual measures (predictor variables, see Table 1), DVA was significantly anticorrelated with NPA (Spearman’s ρ= −0.46, p<10^-4^) with a statistical tendency for anticorrelation with the accommodative facility (ρ = −0.24, p=0.062), both measures being significantly anticorrelated with age and SVA (Fig. 3 and Supplementary Fig. 1). A multiple linear regression on all visual variables explained 28% (p=0.004) of the total DVA variance, with no significant regression slopes for binocular variables when both age and SVA were accounted for, suggesting that age negatively affects the accommodative measures, and these deficits are associated with DVA decline. Vergence measures (associated phoria, NPC, AC/A and Sheard’s ratio) were not associated with DVA (p>0.25).

**Figure 3.**
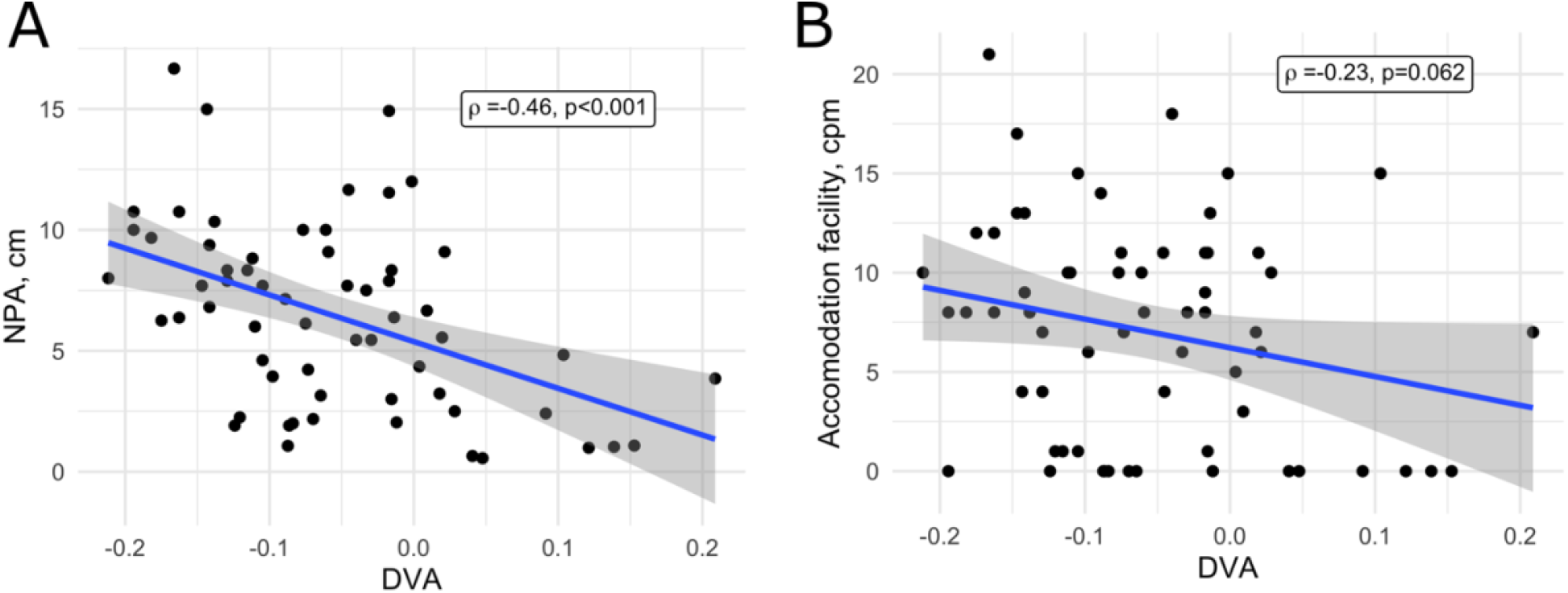
Dependence of the NPA (**A**) and accommodation facility (**B**) on DVA. Each dot corresponds to a participant. Linear regression lines (±*SE*) are shown in blue.

### 3.3. Motor contributions to DVA decline

Here we focused on DVA measured during walking, a condition in which the contribution of motor variables is the strongest. Out of 8 motor variables (see Methods), DVA was significantly anticorrelated with the antero-posterior movement of the smartphone (ρ=- 0.30, p=0.02). As before, it was also correlated with chronological age (ρ=0.48, p<10^-4^) and SVA (ρ=0.57, p<10^-5^), see also Supplementary Fig. 2. Multiple regression analysis revealed that all motor variables together explained 46% of the total variance (F_10,48_=5.858, p<10^-4^). The mean smartphone distance and lateral movement were significant regressors (distance: t= 2.78, p=0.008; lateral movement: t= 0.06, p=0.039), in addition to the antero-posterior movement, age and SVA.

To further explore the role of the smartphone distance, we analyzed it separately for the two DVA measurement methods, since motor-related adaptations are expected to play an important role in the more natural BVA, but not in the AVA (Fig. 4A). Using a two-way ANOVA with the smartphone distance as a dependent variable, age as a between-group factor and DVA mode as a repeated measure, we found a significant main effect of age (F_3,54_ =3.555; p=.02; η^2^ =.165) and of the measurement method (F_1,54_ =70.941; p<10^-10^; η^2^ =.568), as well as a significant interaction (F_3,54_ =7.346; p<0.001; η^2^ =.29). When the BVA method was used, ANP and PO groups held the smartphone at a similar distance (t=-0.02, p=0.99), which was significantly higher than that of the YNP group (YNP vs ANP/PO: t=-2.41, p=0.02) and significantly lower than the MP group (t=-5.42, p=<10^-6^). When the AVA method was used, the MP group was different from all the others (p<0.02) with no other significant differences.

**Figure 4.**
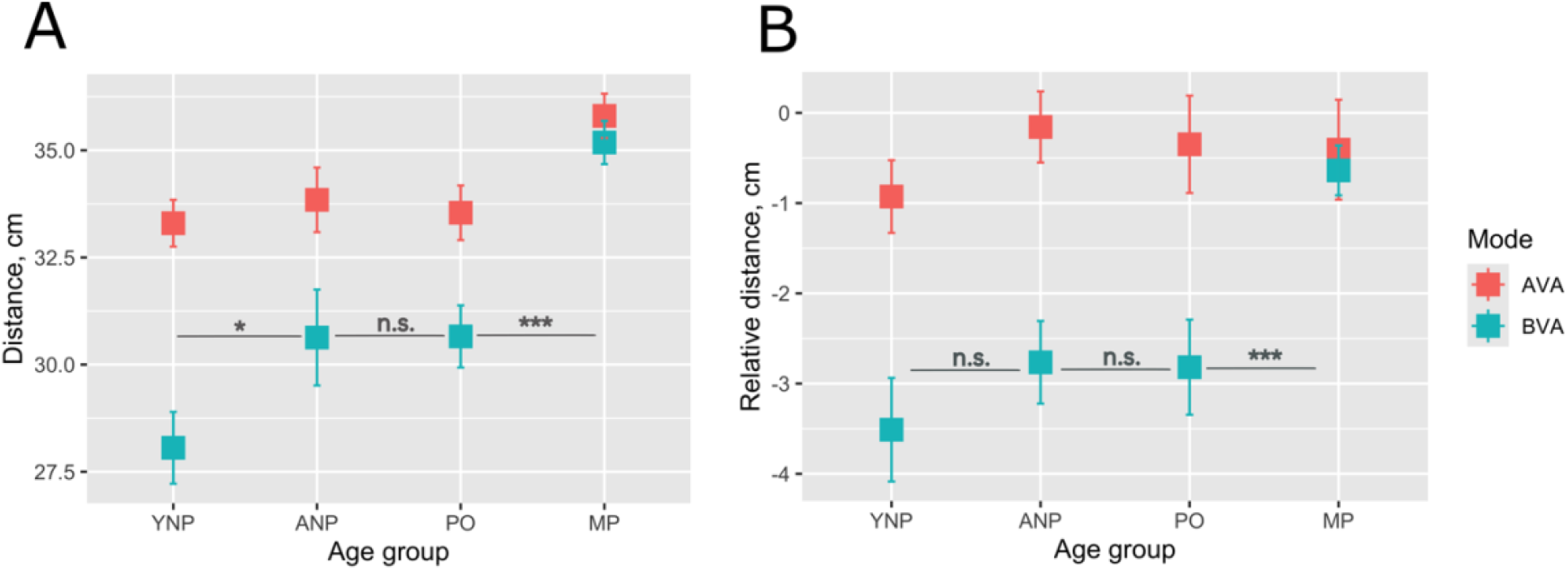
**A**. Smartphone holding distance as a function of age group during DVA measurements performed using AVA and DVA methods. **B**. Relative smartphone distance as a function of age group and measurement method. Larger negative values correspond to a closer holding distance. Significance values are shown only for BVA: *** (p<0.001), * (p<0.05), n. s. (p>0.05).

Finally, the change in holding distance caused by task difficulty was analyzed by a two-way ANOVA with age and DVA mode as factors (Fig. 4B). Both main effects were significant (Age group: F_3,54_=3.538, p=0.02, η^2^ =.164; Method: F_1,54_=24,609, p<10^-5^, η^2^ =.313), such that the MP group was different from all the others in the BVA mode (p<0.01).

## 4. Discussion

Using a new measurement method, imitating a frequently used real-world task, we found an abrupt change in DVA during presbyopia onset (40-46yo). This is in contrast to earlier studies suggesting a more gradual age-related decline in this ability as well as a relative stability of DVA performance until the age of 50 (Chen et al., 2023; Li et al., 2014). A key difference of our study is the possibility to use combined binocular and motor adaptation mechanisms to facilitate retinal image stabilization, in contrast to screen-based tests used in previous studies with middle-aged participant groups. The role of sensorimotor factors beyond purely retinal mechanisms is an essential part of the DVA as a measure of mobility-related performance in dynamic condition such as walking, driving and sports activities (Long & Penn, 1987). While the role of presbyopia in age-related DVA decline may have been reasonably suspected, we are not aware of any previous studies directly assessing it.

While the distance for smartphone holding during walking is approximately 31 cm (Yoon et al., 2019, see also Fig. 4), the maximum accommodation for the age range of the presbyopia onset is about 2.77 diopters (Anderson et al., 2008). This restriction results in a difficulty to focus below a viewing distance of 36.1 cm and increased accommodative requirements (Boccardo, 2021; Mordi & Ciuffreda, 2004), leading to the use of motor compensation strategies. Our data support previous findings (Deshpande et al., 2013; Starkov et al., 2020; Verbecque et al., 2018), showing that DVA decline is strongest during dynamic (e.g. walking on a treadmill) as opposed to static (i.e. sitting and standing) conditions. We note that in our setup, participants were free to choose their walking speed and adjust the smartphone position, whereas in the previous studies in walking participants the speed was controlled by the treadmill and the stimuli were presented on a stationary support (a wall or a computer screen). During our unrestrained walking conditions, the DVA performance of the PO group was statistically indistinguishable from that of the MP group despite a strong age difference (15 years on average). A more detailed analysis of the effect of age on DVA loss (Fig. 2C) shows that the contribution of the dynamical component is temporary, starting during presbyopia onset, reaching its peak near the age of 50 and progressively decreasing afterwards, likely due to long-term sensorimotor adaptations. We note the possibility that PO group consists of two distinct groups of well-adapted and maladapted participants, since 43% of participants in the PO group had DVA loss close to or above the upper limit of the 95%-confidence interval of the regression line, whereas in 35% of participants the DVA loss was close to or below the lower limit.

Our correlation and regression analyses show that the strong deterioration of DVA with age is associated with changes in accommodative mechanisms (NPA and facility of accommodation) as well as in smartphone distance-related measures, with minimal impact of variables related to vergence. The accommodative system has a minimum time required to adjust and refocus on a stimulus that is rapidly moving or changing size in the depth plane (e.g., the accommodation time is 400ms in response to a stimulus shifting by 33 cm, Heron et al., 2001). A slow adjustment to changes in the accommodative demand could thus lead to a decrease in DVA, especially if the movement overpasses the accommodative capacity.

The comparison of the smartphone distance measures (Fig. 4A,B) and DVA dependence on age (Fig. 2B), reveals that PO group has a very particular sensory-motor profile. Indeed, the DVA performance in this group is similar to that of the MP group and different from non-presbyopes, whereas its distance and relative distance measures were statistically identical to the ANP group and very different from MP. Thus, DVA decrease in the PO group can be caused by a lack of behavioral adaptation to the presbyopia onset. This hypothesis is supported by a relatively stronger contribution of motor variables in explaining DVA variability.

Collectively, these results highlight the multifaceted nature of age-related DVA decline, with contributions from both sensory and motor factors, and emphasize the unique challenges faced by individuals undergoing the transition into presbyopia. A survey study of attitudes and beliefs towards presbyopia (Hutchins & Huntjens, 2021) has shown that among participants aged between 36 and 48, only 25% wore a near vision correction, whereas 65% were not aware of presbyopia and its consequences. Our findings suggest that early interventions aimed at promoting awareness of presbyopia and encouraging adaptive behaviors—such as optimal viewing distances and accommodative training—may help mitigate the decline in DVA during this transitional phase. Moreover, the observed heterogeneity within the PO group underscores the importance of personalized approaches in both assessment and intervention. Future longitudinal studies could clarify whether sensorimotor adaptation can be accelerated or guided through specific training protocols, ultimately improving functional vision outcomes during aging.

### 4.1. Conclusion

This study demonstrates that the onset of presbyopia is associated with a sudden and measurable decline in DVA, contrasting with earlier evidence of gradual age-related decline. By employing a more ecologically valid task involving unrestrained walking and smartphone use, we identified significant contributions of accommodative limitations and insufficient behavioral adaptation to the observed performance changes. Our findings emphasize the dynamic and sensorimotor complexity of DVA as a real-world functional measure, particularly during the transitional phase of presbyopia. Given the established impact of presbyopia on quality of life (Goertz et al., 2014) – including reduced productivity, mobility challenges, and overall visual function – these results underscore the importance of early detection, increased public awareness, and timely correction strategies to mitigate visual decline and maintain daily functioning in aging individuals.

## Data Availability

All data produced in the present study are available upon reasonable request to the authors

## 5. Acknowledgements

This work was supported by the French national science association (ANR), LabEx LIFESENSES (ANR-10-LABX-65) and University-Hospital Institutes (IHU) FOReSIGHT (ANR-18-IAHU-01), as well as by the CIFRE ANRt grant (N° 2022/0825).

## Supplementary Material

**Supplementary Figure 1.**
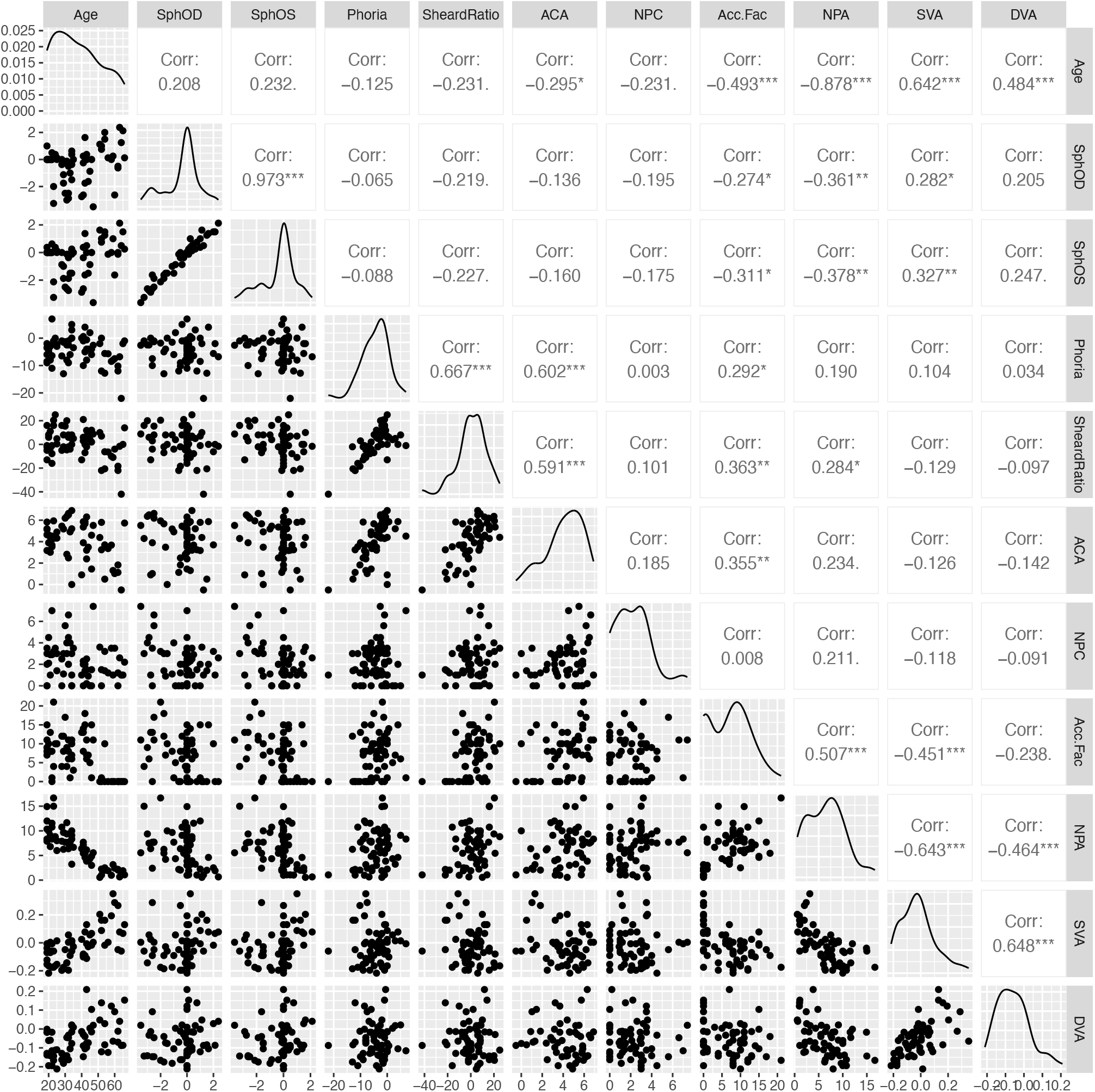
Pairwise correlation plots (lower triangle), approximate distributions (diagonal), and Spearman correlation coefficients between all visual variables, SVA and DVA.

**Supplementary Figure 2.**
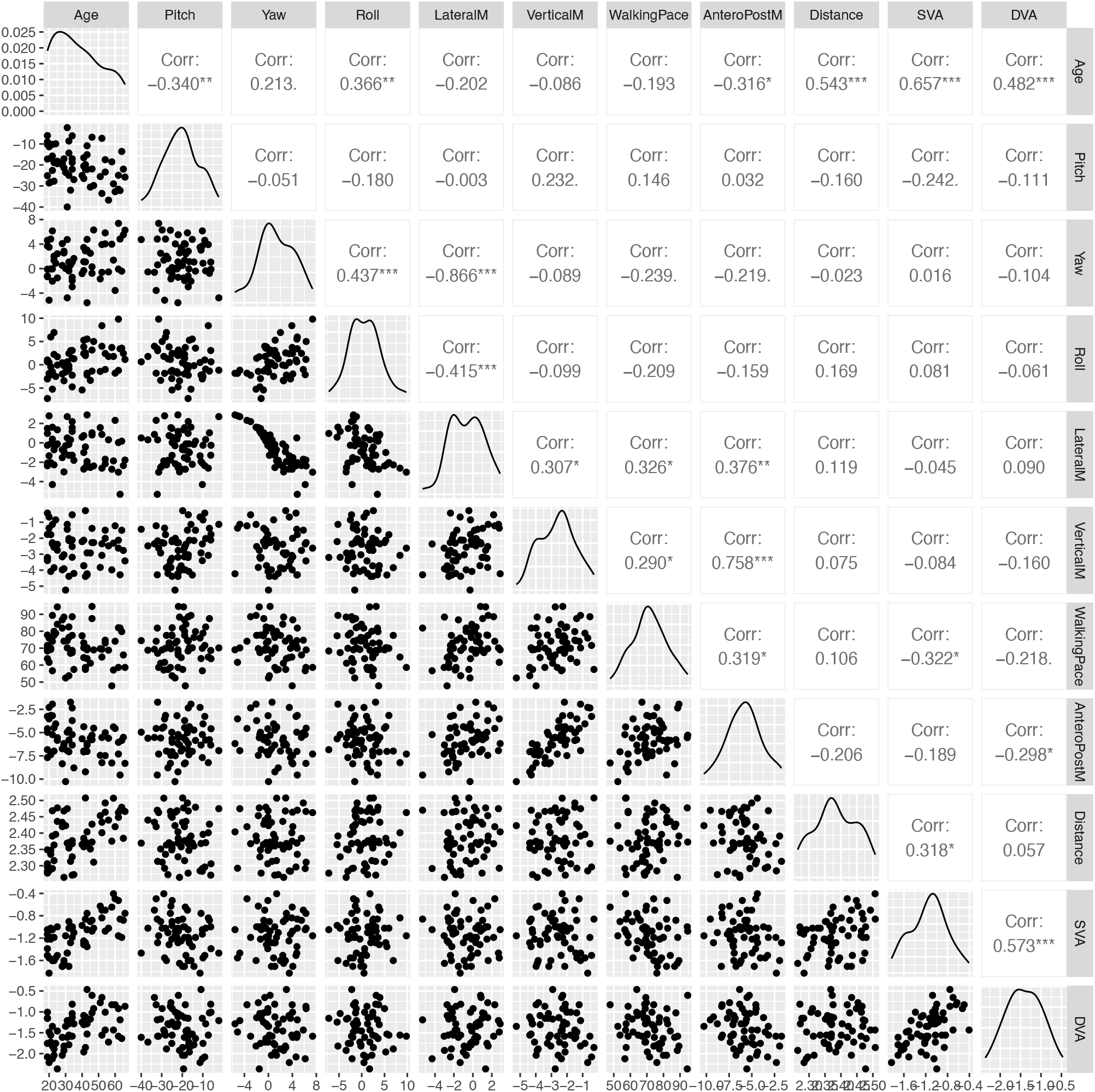
Pairwise correlation plots (lower triangle), approximate distributions (diagonal), and Pearson correlation coefficients between motor variables, SVA and DVA during walking.

